# Time trend analysis of all paediatric suicides in Sweden 1980–2023: Call for awareness and action

**DOI:** 10.1101/2025.11.24.25340902

**Authors:** Sebastian Hökby, Emma Eliasson, Anna Lundh

**Affiliations:** National Centre for Suicide Research and Prevention; Department of Learning, Informatics, Management and Ethics, Karolinska Institutet, Stockholm, Sweden; National Centre for Suicide Research and Prevention; Centre for Health Economics, Informatics and Healthcare Research; Stockholm Health Care Services, Stockholm, Sweden; Section of Psychiatry, Department of Clinical Neuroscience, Karolinska Institutet, Stockholm, Sweden; Centre for Psychiatric Research, Department of Child and Adolescent Psychiatry, Region Stockholm, Sweden

**Keywords:** Suicide, Cause of death, Paediatrics, Child psychiatry, Epidemiology, Public health

## Abstract

**Background:** Current literature on Swedish child suicides relies on data from 2000 and onwards, at most. Presenting four decades of data, this paper aims to advance our understanding of youth suicide patterns. Focusing on the paediatric population, suicide statistics for age groups 10–17 and 10–14 were analysed for time trends.

**Methods:** Data extracted from the Swedish cause-of-death register covered 44 years (1980–2023). Vital records included *n* = 878 suicides in ages 10–17 and *n* = 205 in ages 10–14. Sensitivity analyses additionally included *n* = 152 and *n* = 42 deaths of undetermined intent for each group. Time-series analyses of national suicide rates, stratified by sex, were data-driven and conducted using joinpoint software, estimating Annual Percentage Changes (APC%) and linear splines.

**Results:** Male rates remained stable during the entire period (*P* > 0.163), while female suicides increased linearly (*P* < 0.0001), without joinpoints. The annually increasing female rate was lower in age group 10–17 (APC = 2.33%; CI: 1.35 to 3.31) compared to 10–14 (APC = 4.55%; CI: 3.61 to 6.32). During the first ten years of observation, the male-to-female suicide ratio was 1.9 and 3.4 for the respective groups (10– 17 vs. 10–14). These ratios diminished in the last ten years (1.1 vs 0.8) to become female-dominated in 2023 (0.8 vs 0.9).

**Conclusions:** Female suicide rates in the two age groups increased linearly by over 2% per year. In 2023, male sex was no longer associated with increased risk. Implications for strategic healthcare initiatives are discussed.

**Key messages:** 1. **Research question:** What are the long-term (years 1980–2023) suicide time trends in the Swedish paediatric population, age groups 10*–*17 and 10*–*14?

2. **Findings:** Analysis of *n* = 878 suicides (49% males) indicated stagnant male suicides, while the female suicides in these two age groups increased linearly and annually by 1.6% and 4.5%, respectively – with no joinpoints identified.

3. **Importance:** Information should be strategically disseminated to health care specialists about the increasing female suicide rates, in contrast to stagnant male rates.

## Background

Suicide in childhood and early adolescence is rare but devastating, and its antecedents are often difficult to recognise^1–3^. Detection is further complicated by the fact that a substantial minority of young people who die by suicide have no documented history of ideation, self-harm, mental illness, recent adverse events, or service contact; in a British series of 544 suicides (ages 10–19), roughly 30% fell into this category, and under-recognition or under-reporting of warning signs by families was suspected^4^.

The annual suicide count is small among Swedish youth, aged below 18. Over the past decade (2014– 2023), the average annual count was approximately 24 deaths, of which 8 deaths occurred in the age group 10–14 years. In Sweden, the public debate about child- and adolescent-suicide trends has been constrained by the time-related data range. Published national information has largely relied on data beginning in year 2000^5^ and the Public Health Agency of Sweden typically only reports data from 2006 and onwards^6,7^. Consequently, when inferences are drawn from these data, which predate back to year 2000 or 2006, long-term patterns may become mischaracterized or not reported at all.

Unless sufficiently long time series are utilised, the small suicide counts sometimes make statistical analyses underpowered to detect epidemiologically meaningful subgroup differences, or to identify longitudinally shifting time trends (‘Joinpoints’; a sharp kind of ‘Spline’)^8^. International evidence underscores why the chosen observation period matters. In the United States, paediatric suicide rates, including those of children aged 8–12, have increased during recent decades. Previously, suicide rates in this age group were larger among boys. However, recent findings suggest that female suicide rates have now, in recent years, grown similar to those of males^9^.

To our knowledge, only one prior nation-wide Swedish study has examined paediatric suicide time trends^5^. The study analysed data between 2000–2018 and reported increasing rates among youth below 18 (suicide rate = 1.1 per 100 000, increasing by 2.2% annually). Another finding was that hanging and train/traffic fatalities predominated the suicide methods (79% of cases), while significant sex differences were not found beyond the use of firearms. This is an uncommon method in Sweden^10^. Again, this study was limited by its restricted data scope (19 years) and did not assess the presence of joinpoints (trend-shifts in the time series). Together these drawbacks beg the question of *when* the reported trajectories began, and whether they became mischaracterized by a restricted data period. This study addressed this knowledge gap.

### Aims and objectives

The objective of this study was to present Swedish suicide statistics over an extended 44-year period (1980–2023), focusing closely on two paediatric age groups: 10–17 and 10–14 years. The objective was to do so with high-quality, sex-stratified, cause of death data. We also examined the effect of adding ‘Undetermined intent’ to assess diagnostic bias. The primary aim was to conduct an exploratory (data-driven) investigation of Swedish youth suicides, to estimate the relevant time trends and to identify any joinpoints. The goal was to assess whether the obtained effect results are consistent with the narrative derived earlier, from more restricted studies, starting year 2000.

## Methods

### Ethical considerations and approvals

No data on living humans were analysed. Ethical approval to analyse and report aggregated data findings from the Swedish Cause of Death Registry was obtained from the Swedish Ethical Review Authority (Ref. No. 2023-04783-01), in 29-Oct-2023. The National Board of Social Affairs and Health accepted this ethical decision to use the data for research purposes in conjunction with data delivery (Ref. No. 122437/2023), on 26-Sep-2024. The Swedish Ethical Review Authority further granted an extended ethical approval (Ref. No. 2024-07193-02) on 29-Dec-2024 to share aggregated registry data with international research partners and scientific journals, to contribute to global research collaborations. Although the death count was sometimes small, the data are not connected to any other personally sensitive information that can be connected to personal identifiers (e.g., socio-economic variables or local living areas). The study adhered ethically to the Declaration of Helsinki, and to the RECORD guidelines for reporting. The aggregated data and its metadata are shared openly^11^.

### Data description

The Swedish National Centre for Suicide Research and Prevention (NASP) held relevant cause-of-death data for the entire 44-year period (1980*–*2023), provided by the Swedish National Board of Social Affairs and Health, in 2024^12,13^. Extraction included all Swedish children aged <18 years during the 44-year period. Data of primary interest included death counts, or rates, with determined suicidal intent (Intentional self-harm, corresponding to ICD-9 and ICD-10 diagnoses E950*–*E959, and X60*–*X84, respectively). Deaths with undetermined intent were only included in bias sensitivity analyses (ICD-9 = E980*–*E989; ICD-10 = Y10*–*Y34).

The raw dataset^11^ contains the annual counts of each diagnosis type (X60–X84 vs X60–X84 + Y10– Y34), stratified by sex, but aggregated by year and age group. ‘Sex’ refers to binary gender assigned at birth. Missing data on exact day of death were permitted, as only annual data were needed. To calculate crude suicide rates, the annual suicide count was divided by its mid-year population count, multiplied by 100 000. Thus, a given mid-year population count (e.g. 1980) was calculated by averaging the end-year count for two consecutive years (e.g. 1979 + 1980 ÷ 2). The average annual population count during the study period was *M* = 887 403 (SD = 62 744) in age group 10*–*17, and *M* = 550 930 (SD = 47 548) in age group 10*–*14. Population counts were accessed from Statistics Sweden^14^. Mid-year counts are included in the raw dataset.

The final dataset comprised of *n* = 1030 cases (deaths) in the age group 10*–*17, of which *n* = 603 (59%) were males and *n* = 427 (41%) were females. These numbers include *n* = 98 (16%) male cases, and *n* = 54 (13%) female cases of death with undetermined intent. Regarding age group 10*–*14, a total of *N* = 247 cases were available for analysis, of which *n* = 133 (54%) were males, and *n* = 114 (46%) females. Included are *n* = 27 (20%) male and *n* = 15 (13%) female cases of death with undetermined intent. Deaths in age group 0–9 were all excluded, as only *n* = 3 suicides were observed, and all other deaths had been classified with undetermined intent.

### Statistical modelling

Joinpoint regression software (v.5.4.0)^15^ was used to estimate time trends and joinpoints. It has previously been used in suicidological research^16^. The effect size measure of interest was the Annual Percentage Change (APC%), including its 95% confidence interval (CI), calculated using the empirical quantile method^17^. Any replication attempt ought to utilise this specific version to ensure result correctness, and models should be defined using Sex as a stratifying “By-Variable” (categorical joinpoint variable). Model fit was assessed by comparing the data-driven Weighted Bayesian Information Criterion (WBIC: “smaller is better”). WBIC was used in favour of the computationally heavy permutation test method, but results could be replicated with the permutation method^17^. Permutations may produce more robust parameter estimates and joinpoints, but WBIC performs better in small sample sizes, and more sensitive to finding them when counts are small. Using 44 annual datapoints maximizes the degrees of freedom to identify seven joinpoints, with minimum two data points in between each joinpoint^18^. This is also the default joinpoint setting (v.5.4.0). Regarding the age group 10*–*17, the crude suicide rates between 1980*–*2023 were modelled after they had been calculated *within* the joinpoint software to automatically produce heteroscedastic standard errors^19(p2)^. However, the homoscedastic model outperformed the corresponding heteroscedastic model as the latter exhibited negative autocorrelation (male vs female: AR^1^ = −0.21 vs −0.46). We thus decided on the homoscedastic model (constant variance) error modelling of suicide rates. Logarithmic data transformation with identity-link was used to model suicide rates, as counts rates per 100 000 capita The assumption of uncorrelated errors in vital record data analysis is common practice according to the time trend guidelines by the U.S. National Center for Health Statistics^18^.

Regarding age group 10*–*14, the final selected model used homoscedastic Poisson error variance to analyse the trend in suicide counts, unadjusted for population size^19^. The homoscedasticity option precludes the use of an autocorrelation function. The dataset contained several cells with value = 0, especially between years 1980*–*1985. In such instances, the joinpoint software imputed Count = 0.5 into empty cells, before log-transforming and running the analysis (zeros cannot be modelled). Note that straight slopes appear to be curved due to log-transformation.

## Results

The main result and time trend for age group 10*–*17 is illustrated in *Figure 1*. The female suicide rate increased steadily in a linear fashion, without any joinpoints, over the 44-year-period (APC = 2.33%; CI: 1.35 to 3.31; *P* < 0.0001). The male slope visualised in *Figure 1* shows no meaningful incline or decline and is not significantly different from zero (APC = 0.48%; CI: −0.48 to 1.47; *P* = 0.3299). Although male and female CIs overlap, suicide rates became more female-dominated over time. Sensitivity analysis showed that results held true when also including undetermined cases, although this produced weaker effect sizes (*male* APC = −0.10%; CI: −1.11 to 0.90; *P* = 0.8226; *female* APC = 1.64%; CI: 0.67 to 2.61; *P = 0*.*0020*). Only linear trends, but no joinpoints (linear splines) were indicated, regardless of sex.

**Figure 1.**
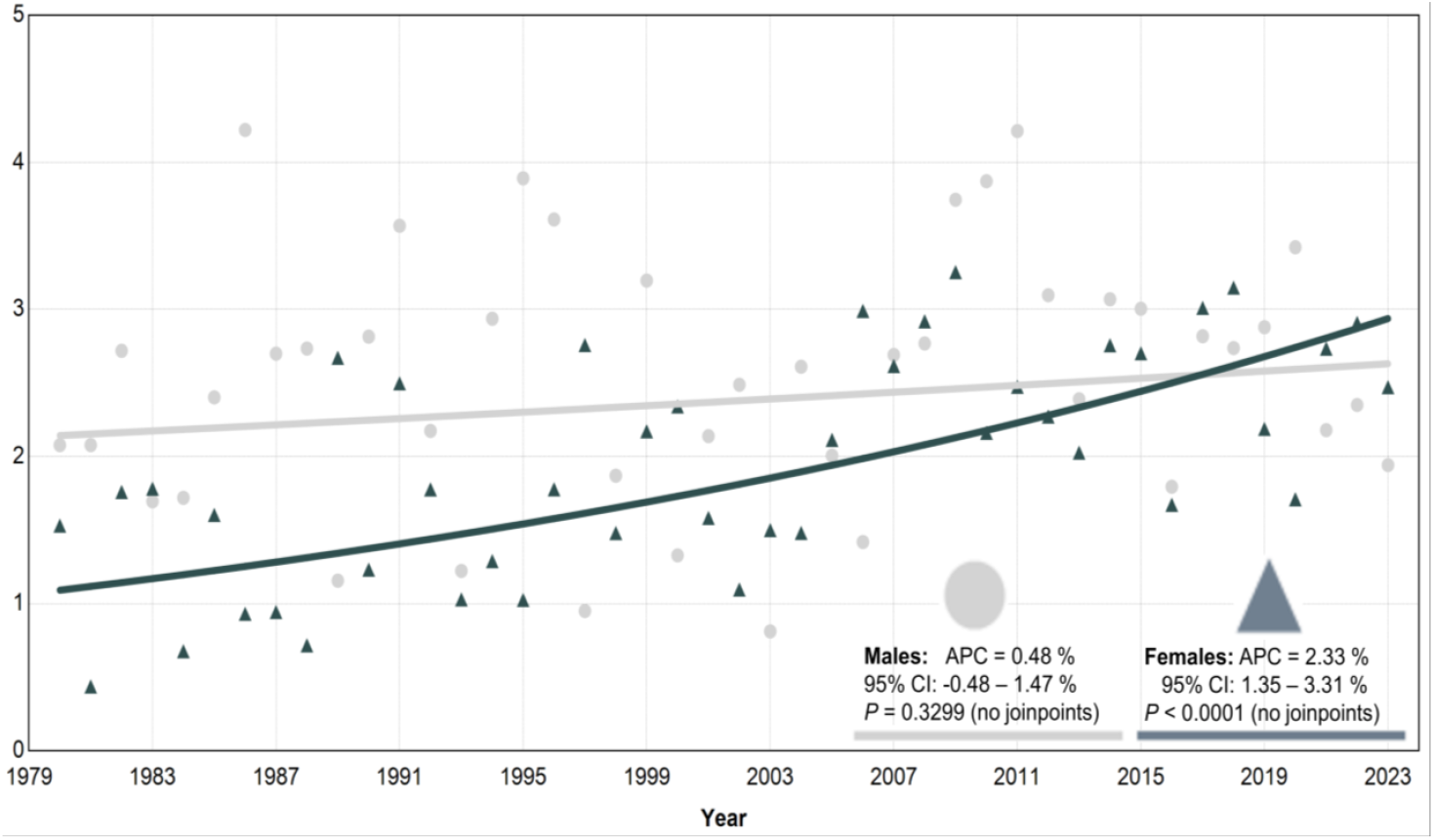
Age group 10–17: *S*uicide rates (per 100 000) over 44 years. APC = Annual Percentage Change.

The main result and time trend for age group 10–14 is illustrated in *Figure 2*. The female suicide count increased significantly in a linear fashion, without any joinpoints, over the 44-year-period (APC = 4.55%; CI: 3.61 to 6.32; *p* < 0.0001). The male slope was virtually horizontal and not significantly different from zero (APC = 0.83%; CI: −0.38 to 2.30; *P* = 0.1628). Male and female CIs do not overlap, and suicide counts became increasingly female-dominated over time. Sensitivity analysis showed that results held true when also including undetermined cases but produced weaker effect sizes (*male* APC = 0.12%; CI: −1.06 to 1.40; *P* = 0.7914; *female* APC = 4.16%; CI: 3.10 to 6.08; *P* < 0.0001).

**Figure 2.**
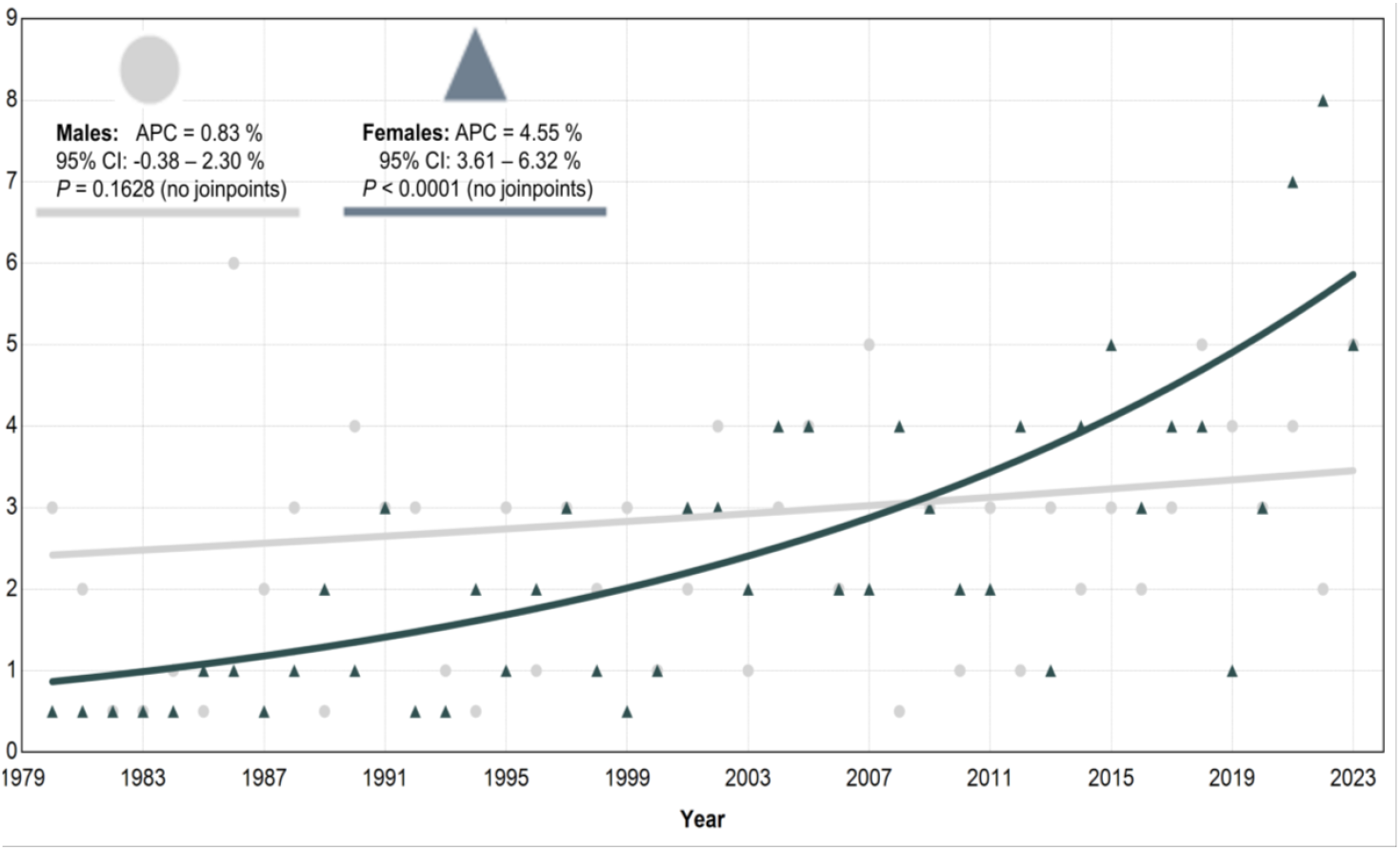
Age group 10–14: Suicide counts over 44 years. APC = Annual Percentage Change.

Specific suicide rates and male-to-female ratios can be derived from the shared raw dataset^11^. For instance, during the initial ten years (1980*–*1989), the male-to-female suicide ratio was 1.9 and 3.4 in the respective groups (Ages 10–17 vs Ages 10–14). In the last ten observation years (2014*–*2023), these ratios had diminished to 1.1 and 0.8. In year 2023, both age groups exhibited a female-dominated suicide ratio (0.8 and 0.9), unlike any other age group (*Table 1*).

**Table 1.** Swedish suicide rates ^a^ (per 100 000) in various age groups, year 2023 ^b^.

| Age group | Both sexes | Male | Female | Male/Female ratio <sup>c</sup> |
| --- | --- | --- | --- | --- |
| Ages 0–4 | 0 | 0 | 0 | n/a |
| Ages 5–9 | 0 | 0 | 0 | n/a |
| **Ages 10–14** | **1.58** | **1.54** | **1.63** | **0.945** |
| **Ages 10–17** | **2.20** | **1.94** | **2.47** | **0.786** |
| Ages 15–19 | 6.58 | 7.98 | 5.09 | 1.568 |
| Ages 20–24 | 13.25 | 16.74 | 9.35 | 1.790 |
| Ages 25–29 | 13.60 | 18.07 | 8.83 | 2.046 |
| Ages 30–34 | 13.97 | 18.73 | 8.95 | 2.093 |
| Ages 35–39 | 15.31 | 23.72 | 6.42 | 3.695 |
| Ages 40–44 | 13.35 | 18.59 | 7.86 | 2.365 |
| Ages 45–49 | 12.80 | 17.27 | 8.16 | 2.116 |
| Ages 50–54 | 16.20 | 21.23 | 11.04 | 1.923 |
| Ages 55–59 | 18.95 | 27.26 | 10.42 | 2.616 |
| Ages 60–64 | 18.76 | 26.71 | 10.72 | 2.492 |
| Ages 65–69 | 16.02 | 23.76 | 8.40 | 2.829 |
| Ages 70–74 | 16.36 | 22.53 | 10.51 | 2.144 |
| Ages 75–79 | 15.56 | 24.59 | 7.27 | 3.382 |
| Ages 80–84 | 17.57 | 28.75 | 8.11 | 3.545 |
| Ages 85–89 | 27.87 | 54.42 | 9.60 | 5.669 |
| Ages 90–94 | 20.36 | 45.67 | 7.64 | 5.978 |
| Ages 95+ | 33.66 | 70.08 | 22.15 | 3.164 |
| **All ages (0-95+)** | **12.60** | **17.77** | **7.36** | **2.414** |

<sup>a</sup> Suicides are defined by diagnosis codes X60–X84 in the tenth revision of the *International Statistical Classification of Diseases and Related Health Problems* (ICD-10).
<sup>b</sup> Rates and ratios for years 1980–2022 can be calculated from the raw dataset<sup>11</sup>.
<sup>c</sup> Male/Female ratio = Dividing the male rate by the female rate. The age spans 10–14 and 10–17 are especially highlighted as they are the centre of this study. These two groups exhibit an “inverse gender paradox in suicide”, unlike any other group.

## Discussion

This study examined all suicides in under-aged Swedish children over a 44-year period, analysing time trends in official cause-of-death data (1980–2023), using joinpoint software. Focusing on two paediatric age groups, 10–17 and 10–14, it was clear that the female rates increased while the male rates were stable. The study shows that previously reported Swedish time trends and patterns date back to 1980; not just year 2000^5^. Thus, a new picture of Swedish paediatric suicides seems to have emerged, with implications for clinical practice and suicide prevention strategies, tailored to at-risk children. Notably, the female suicide rates in the two age groups increased linearly by over 2% per year. The annual percentage change (APC%) was 2.33% (1.35–3.31) in ages 10–17 and 4.55% (3.61–6.32) among girls aged 10–14. The absence of joinpoints indicated a persistent rise rather than episodic surges. Conclusively, male sex has been, but is no longer, a suicide risk factor among Swedish children. Analysis of *n* = 878 suicides (49% males) indicated no change in male suicide rates over the 44 years.

Separating the 10–14 age group seemed clinically and epidemiologically important. Over 44 years, we found a steady annual increases among females, most pronounced in ages 10–14. Annual counts were small in this group, particularly in the early 1980s and 1990s. Thus, reducing the observation period or refraining from sex or age-data stratifications would unlikely have produced as straightforward results. Stratification unveiled changing effect sizes across specific female age groups. By reporting suicide trends separately for ages 10–14 and 10–17, and by extending the observation window to 1980–2023, this study provides an updated view of paediatric suicide in Sweden compared to previous publications, with starting years around 2000 or 2006^5–7^. This study found male slopes which were virtually horizontal, and stagnant across all years, and the result patterns held true in sensitivity analyses which adjusted for deaths with undetermined intent.

Single-year contrasts should be interpreted cautiously, but here, the direction and stability of the long-term trends suggest a meaningful reconfiguration of risk within childhood. Our results replicate and extend the finding of increasing Swedish paediatric suicides previously reported for years 2000– 2018^5^. Trajectories also align with international findings of increasing female suicide rates in ages 8– 12, during years 2001–2022, and a narrowing traditional male predominance in that younger band^9^. The cross-national similarity suggests that broadly acting, time-varying risk environments might be at play (such as psychosocial, digital, familial, school-based, or health service factors). For instance, while the present study did not examine any underlying causes, the iPhone release in 2007 has been hypothesised to increase teenagers’ ‘fear of missing out’, which is a proximal psychosocial exclusion stressor^20^ believed to epigenetically increase the suicide risk, particularly among females^21–24^, but also according to individual differences in social capital^25^.

### Methodological strengths and limitations

Analyses demonstrated a benefit of carefully stratified data^18^. If not done, they might understate the robustness and effect size of age-dependent suicide trends. No data analysis, unstratified by sex, would have generated the present findings, which show that the female suicide increase is larger and more persistent than suggested by shorter time series studies. Observing a long-term linear rise, rather than a recent acceleration, the study helps to explain why shorter study designs might have understated the cumulative change in girls. While boys were over-represented in the 1980s and 1990s (*Figure 1* and *Figure 2*), the 2023 female rates in were greater than, or comparable to, the male rates; a pattern not seen in any other Swedish age group (*Table 1*). Our analyses did not identify any joinpoints that convolute these straight-forward findings, but a study limitation is that our data could not be stratified further, to investigate even more detailed research questions, for example suicide methods^5,10^. Our extended time-series analyses required data imputations for ages 10–14, given the small annual counts in this group. As is true for many registry-based studies, diagnostic classification issues may introduce bias; however, the inclusion of undetermined intent and the absence of joinpoints speaks against a conclusion being driven solely by coding artefacts. We did not model the transition between editions of the International Classification of Diseases, that is, Sweden’s transition to ICD-10 in 1997. Nevertheless, 44 annual datapoints should sufficiently power analyses to detect joinpoints for any reason^18^. Both the WBIC criterion and permutation method produced similar results, but the primary use of WBIC is especially efficient when modelling small numbers^17^. Future studies could further evaluate this methodological trade-off. We next conclude this paper by discussing some practical implications.

#### I. Implications for clinical practice and training

Primary care and child psychiatry should embed age-attuned suicide risk assessment into routine care, including for pre-adolescents. Safety planning must involve both children and caregivers, and clinicians should be trained to recognise developmentally specific presentations (e.g. indirect or metaphorical expressions of hopelessness or death themes)^4,26,27^. A modernised conceptual framework should acknowledge that a purely chronological view of child development is insufficient. Suicide prevention/postvention should adopt a developmental perspective that foregrounds biological, psychological, and social maturity, recognising that suicidal ideation and behaviours in children may not be verbally explicit^26,28^.

#### II. Implications for Swedish child and adolescent psychiatry

Professionals should restate that the typical male excess in suicide mortality seen at older ages does not apply in the same way to children in recent years; female sex appears more salient in current Swedish child suicide mortality patterns, particularly ages 10–14, acknowledging that sex is not modifiable and should be contextualised within comprehensive risk formulations^29^. Waiting until mid-adolescence may miss critical windows for a subset of at-risk children.

#### III. Implications for schools and social services

Risk assessments in child protection should integrate the elevated salience of suicidality in younger children, particularly amid family adversity and parental mental illness/substance use. Family-inclusive interventions are needed to strengthen proximal protective factors^4^. Schools should be equipped with rapid referral pathways for suicidal students, have postvention routines, and staff trained to observe suicide warning signs.

## Conclusion

This is the first study to show that in female paediatric suicide rates have increased in Sweden over four decades, in contrast to stagnant rates seen among males. It calls for paediatric suicide to be treated as a growing public health concern, requiring coordinated action among healthcare professionals, social service workers and school staff. It also calls for further research to map the most important mechanisms driving these time trends. Such information is pivotal to guiding the development of broad paediatric suicide prevention strategies, as well as clinically targeted interventions.

## Data availability

The study dataset is openly accessible via DOI: https://doi.org/10.48723/1546-qm94 License: Public Domain Marked 1.0 (published online: 24th November 2025).

## Author statements

No author has any Conflict of Interest or Funding to declare.

